# PRECISE: Benchmarking digital pathology with expert-annotated contiguous IHC–H&E serial prostate sections

**DOI:** 10.64898/2026.07.21.26358559

**Authors:** Adriana K. Calapaquí Terán, Abel A. González Bernad, Miriam Cobo Cano, Lucía Sánchez Magdaleno, Sara Marcos González, Roberto Carlos Delgado Bolton, Jaled Moustafá Calvo, José Javier Gómez Román, Lara Lloret Iglesias

## Abstract

We present PRECISE (PRostate Expert-annotated Contiguous IHC–H&E Serial sEctions), a hybrid histopathology dataset of paired hematoxylin and eosin (H&E) and immunohistochemistry (IHC) whole-slide images (WSIs), comprising 37 prostate core needle biopsies from 25 patients, each with matched H&E and CKAPM+racemase staining. To the best of our knowledge, this is the first publicly available dataset offering spatially harmonized, pixel-level expert annotations across both staining modalities in prostate biopsy WSIs — directly mirroring the two-stage (H&E-then-IHC) clinical diagnostic workflow used to resolve morphological uncertainty, restricted to cases in which that workflow reached diagnostic consensus. The dataset contains 24,387 annotations spanning seven diagnostically critical classes: malignant glands, benign glands, stromal tissue, intraductal carcinoma (IDC-P), high-grade prostatic intraepithelial neoplasia (HGPIN), atypical intraductal proliferation (AIP), and tissue artifacts. Unlike existing resources, which focus on binary tumor classification or lack IHC pairing, this dataset captures the full morphological spectrum encountered in routine prostate pathology, including rare precursor lesions and confounding entities underrepresented in current benchmarks. Annotations were validated through a structured three-stage consensus by two expert uropathologists, with IHC serving as biological ground truth for boundary definition. PRECISE is designed as a robust benchmark for multimodal semantic segmentation and self-supervised learning, and is openly released to promote reproducible research and accelerate AI-assisted diagnosis in prostate cancer. Our dataset is available at 10.5281/zenodo.20721779.

## 1. Background

### 1.1 Scientific and Clinical Motivation

Prostate cancer remains one of the leading causes of oncological morbidity and mortality worldwide. The definitive diagnosis, risk stratification, and subsequent therapeutic management rely heavily on the histopathological evaluation of tissue biopsies and radical prostatectomy specimens. The Gleason classification system, which evaluates the architectural patterns of the prostate glands, represents the gold standard clinically for assessing aggression. However, digital pathology workflows face immense challenges due to the high intra- and inter-observer variability among pathologists, particularly when distinguishing complex or borderline morphological features such as High-Grade Prostatic Intraepithelial Neoplasia (HGPIN), atypical intraductal proliferations, and early-stage intraductal carcinomas. Computer-Aided Diagnosis (CAD) systems driven by deep learning offer a promising avenue to standardize slide analysis, reduce diagnostic turnaround times, and uncover sub-visual computational phenotypes that correlate with patient outcomes.

### 1.2 Identified Gap in Existing Resources

Despite the rapid evolution of computational pathology, a significant bottleneck persists in the development of robust, generalizable machine learning (ML) models for prostate cancer. Deep learning (DL) architectures inherently rely on massive volumes of pixel-level or patch-level annotations curated by expert pathologists. In digital pathology, this annotation process is exceptionally labor-intensive, time-consuming, and cost-prohibitive. Existing public repositories, while valuable, suffer from major constraints. Many datasets are highly homogeneous, originating from single-center cohorts utilizing a solitary whole-slide scanner model, which leads to significant performance degradation when deployed on external, unseen multi-site data due to domain shift. Furthermore, current datasets primarily focus on binary classifications (tumor vs. benign) or localized Gleason score categorization, largely ignoring critical adjacent microenvironmental and confounding histological entities such as stromal variations, specific tissue artifacts, and precursor lesions.

### 1.3 Similar Datasets and Tools

Several benchmark datasets have shaped the current landscape of computational prostate pathology. The Prostate cAncer graDing Assessment (PANDA) challenge dataset represents a landmark resource, providing thousands of whole-slide images (WSIs) with slide-level Gleason scores and pixel-level tissue masks (Bulten et al., 2022). However, PANDA’s annotations are coarse, heavily automated via proprietary algorithms, and lack the granularity required to identify localized sub-visual patterns or atypical precursor entities. Other repositories, such as The Cancer Genome Atlas (TCGA-PRAD) (Zuley et al., 2016), offer rich multi-omic clinical correlations alongside high-resolution diagnostic slides, but they do not provide detailed, regional expert annotations for non-tumor structures, stroma, or benign mimics.

Furthermore, related public datasets explore specific histological representations but differ substantially from our clinical context, immunohistochemistry (IHC) marker selection, annotation methodology, and overarching histological scope. For instance, while the Gleason2019 challenge dataset (Nir et al., 2018) and the Tan Tock Seng Hospital repository (Oner et al., 2022) offer fine-grained glandular annotations, they are constrained by limited categorical scopes or small sample sizes. Even recent extensive cohorts like DiagSet (Koziarski et al., 2024), despite providing millions of multi-pathologist labeled patches, remain restricted to single-modality morphological representations on H&E. Our previously published dataset DOI: 10.5281/zenodo.18930300 addressed some of these existing gaps by providing high-resolution Hematoxylin and Eosin (H&E) whole-slide images of prostate CNBs with detailed pixel-level annotations spanning 28 distinct histological classes, validated by two expert uropathologists using IHC as a reference standard. However, that prior resource remained restricted to single-modality morphological representations, neglecting the multimodal workflow essential for definitive clinical stratification in ambiguous cases.

### 1.4 Statement of Novelty

To address these critical gaps, the present work significantly extends our prior resource 10.5281/zenodo.18930300 by introducing a paired, complementary IHC component stained with the CKAPM+racemase cocktail, establishing a unique hybrid H&E–IHC digital pathology dataset. The distinct novelty of this resource lies in its structured integration of diverse expert-annotated histological regions where both modalities share semantically harmonized, pixel-level annotations across seven highly critical diagnostic classes: tumor, benign glands, HGPIN, intraductal carcinoma, atypical intraductal proliferation, artifacts, and stroma. Here, semantically harmonized denotes that corresponding tissue regions across the two serial sections carry consistent expert-assigned class labels; because H&E and IHC sections are physically distinct, serially cut slices, exact pixel-to-pixel geometric registration between the two modalities is not guaranteed, a constraint discussed further in Section 3 (Known limitations). To the best of our knowledge, PRECISE (PRostate Expert-annotated Contiguous IHC–H&E Serial sEctions) represents the first publicly available hybrid H&E–IHC dataset derived from prostate core needle biopsies featuring expert pixel-level annotations semantically harmonized across both staining modalities. No existing public resource offers paired biopsy-level WSIs with this specific IHC cocktail and semantically consistent expert annotations covering the full spectrum of diagnostically relevant glandular lesions. By providing this dual-modality framework, this dataset is uniquely designed to serve as a foundation for multimodal AI systems that directly replicate the classic two-stage (H&E-then-IHC) diagnostic workflow used in routine clinical practice to resolve diagnostic uncertainty.

## 2. Summary

### 2.1 Vision and Objectives

The ultimate goal of PRECISE is to democratize access to high-fidelity, expertly curated digital pathology data, shifting the paradigm of prostate cancer CAD from simple tumor detection to comprehensive, multi-class microenvironmental parsing. The primary objective is to provide a standardized, robust benchmarking framework for semantic segmentation architectures, panoptic segmentation models, and self-supervised learning representations. By offering a meticulously harmonized set of diagnostic classes, this dataset aims to foster the development of AI algorithms capable of assisting pathologists in routine clinical screening, minimizing diagnostic discordance, and automating the quantitative assessment of complex spatial relationships between benign tissue, precursor lesions, and invasive carcinomas. Figure 1 depicts an example extracted from the dataset showing two whole-slide images (WSIs), one stained with H&E and the other with a immunohistochemical stain, together with their corresponding segmentation masks. Table 1 contains a summary of the number of manually annotated regions per histological class in H&E and HMWCK/AMACR stainings.

**Table 1:**
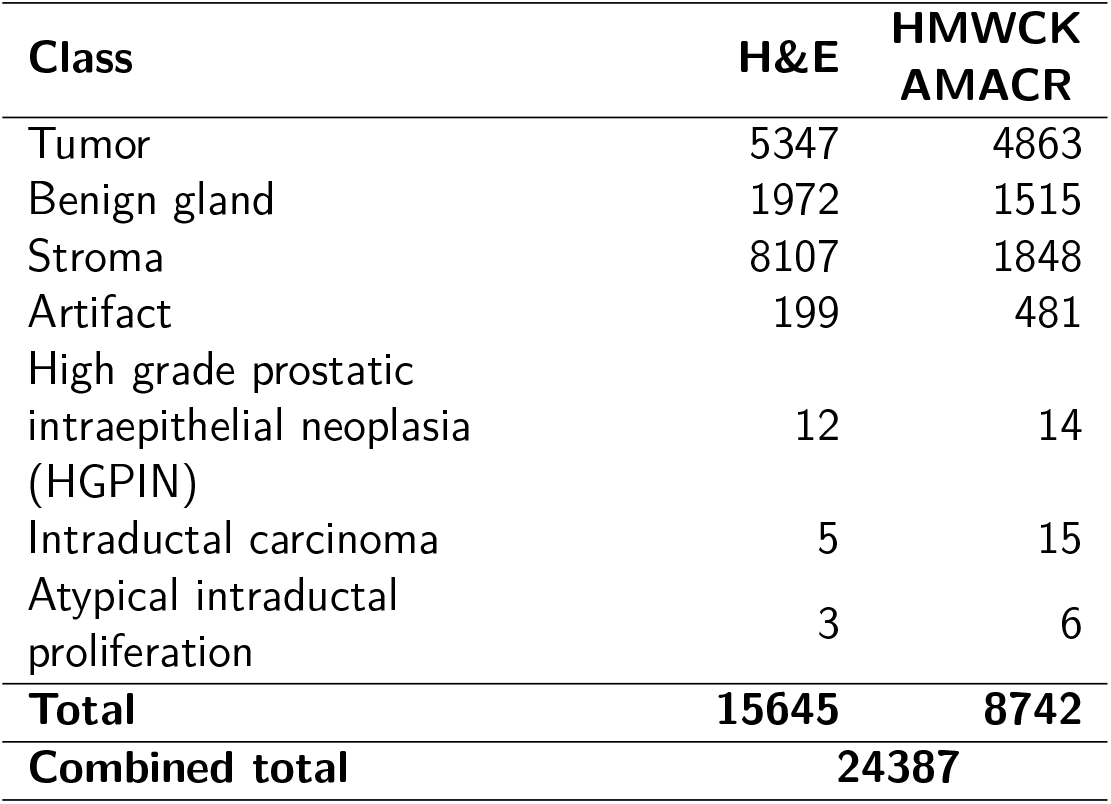
Number of annotated regions (instances) per histological class in H&E and HMWCK/AMACR stainings. All classes were manually annotated except stroma, which was generated through an automated thresholding and morphological-refinement pipeline (see Section 5.3.2).

**Figure 1.**
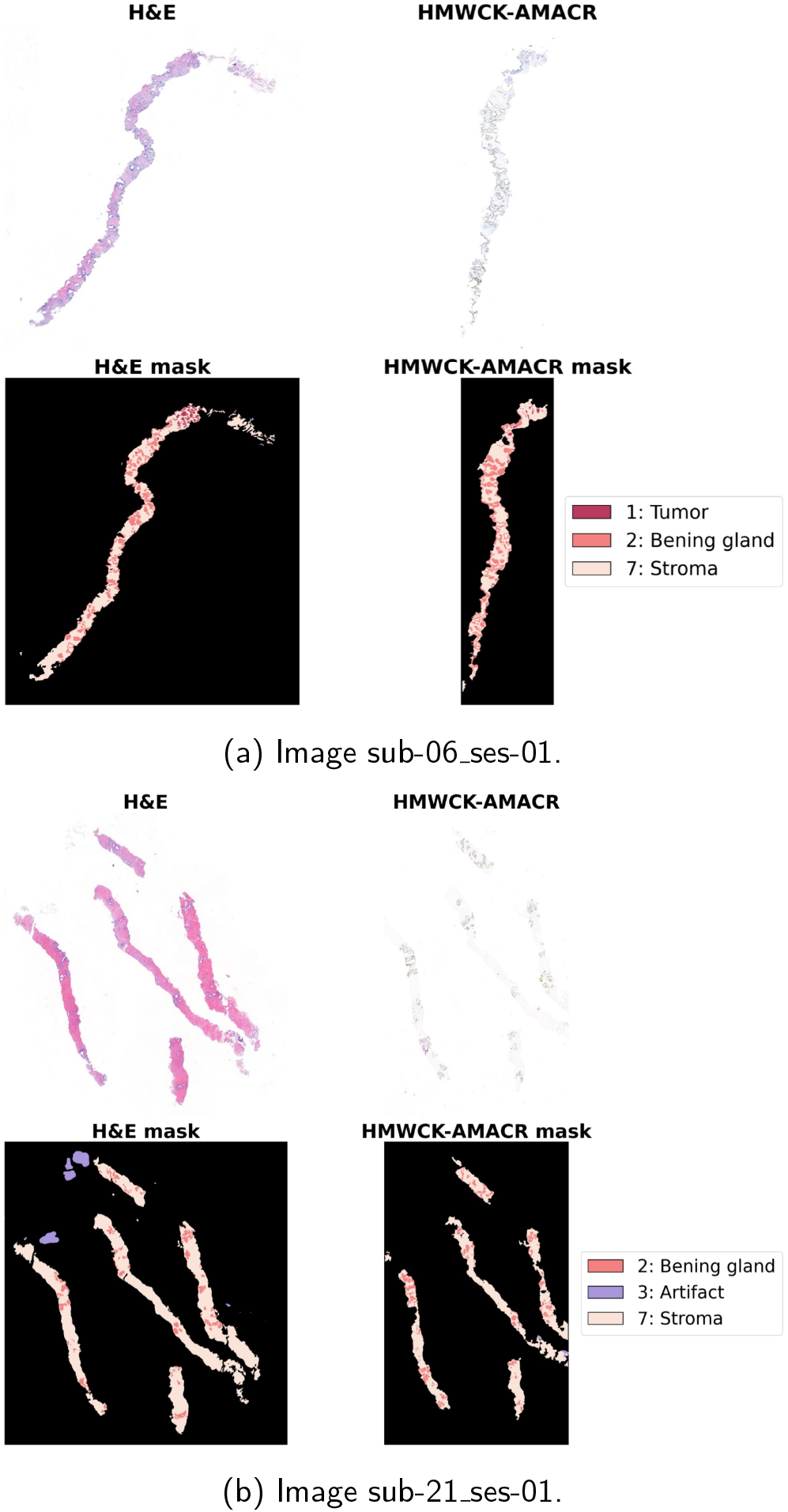
Examples of two whole-slide images (WSIs), one stained with H&E and the other with a immunohistochemical stain, together with their corresponding segmentation masks. Clinical metadata are also provided in the titles for reference.

### 2.2 AI-Readiness Statement

The dataset is ready to be directly integrated in DL pipelines without additional manual preprocessing. More details are given in the Methods section.

### 2.3 Resources Needed to Use the Dataset

To effectively utilize this resource for ML experimentation, users should meet the following technical requirements:

- **Hardware Infrastructure:** Due to the high spatial resolution of the digital pathology data, a dedicated workstation or cluster node equipped with at least one modern NVIDIA GPU (minimum 16 GB VRAM, e.g., RTX 4080, A100, or equivalent) is highly recommended for efficient model training and inference. At least 60 GB of available disk storage is recommended for dataset handling, preprocessing, caching, and model outputs, preferably on a high-speed solid-state drive (SSD) to reduce data loading times and improve overall pipeline performance. The dataset is distributed as pyramidal TIFF WSIs, enabling efficient multi-resolution access and region-based reading without loading the complete slide into memory. This allows visualization and patch extraction even on systems with modest memory resources. However, full-resolution visualization and processing can be memory intensive, and at least 32 GB of system RAM is recommended. A single WSI may occupy up to 7 GB when decompressed as a uint8 array, excluding the corresponding segmentation mask, complementary immunohistochemistry-stained image, and any additional intermediate copies generated during custom analysis workflows.
- **Software Environment:** A standard Python environment is required (version 3.10 or higher). The provided code relies on widely adopted scientific computing and image-processing libraries, including NumPy and SciPy for numerical operations, TIFFFile Gohlke (2022) and Zarr Zarr Developers (2020) for efficient access to pyramidal whole-slide images, and Pillow and Matplotlib for image visualization and exploratory analysis.
- **Data Access Requirements:** No specialized proprietary software or licensing keys are needed to read, visualize, or process the underlying image data and annotation masks.

## 3. Discussion

This work presents a hybrid H&E–IHC dataset of prostate core needle biopsies with pixel-resolution annotations, harmonized at the semantic/class level across serial sections, designed to support the development of multimodal AI systems for computational pathology. Below we critically reflect on its strengths, limitations, representational concerns, and recommendations for responsible use.

### Strengths

The primary strength of PRECISE lies in its hybrid multimodal structure: to the best of our knowledge, no existing public dataset provides paired, serial-section H&E and CKAPM+racemase IHC whole-slide images from prostate CNBs with expert pixel-level annotations harmonized at the semantic level across both modalities. This design directly mirrors the two-stage diagnostic workflow used in routine clinical practice, where IHC is requested as an ancillary tool when H&E morphology alone is inconclusive, and therefore provides a uniquely realistic foundation for training and benchmarking multimodal AI models. Annotation quality is a further strength: all labels were generated by two experienced uropathologists through a structured consensus process, with immunohistochemistry used as a biological ground truth to define tissue boundaries with precision. The inclusion of diagnostically challenging and clinically relevant classes — HGPIN, intraductal carcinoma, and atypical intraductal proliferation — alongside with the more common categories addresses a gap that persists in most existing prostate pathology datasets, which tend to focus exclusively on tumor versus benign discrimination. Targeting HGPIN is particularly valuable from a computational standpoint; prior biological evidence indicates that AMACR expression rates and staining intensity in HGPIN are significantly upregulated when these precursor lesions are adjacent to invasive carcinoma (Wu et al., 2004). By providing paired, semantically harmonized, pixel-level H&E– IHC representations of these regions, our dataset uniquely enables AI models to move beyond binary tumor detection and explore subtle microenvironmental and spatial patterns associated with disease proximity. The annotation harmonization between modalities ensures that the same tissue regions carry consistent class labels in both H&E and IHC images, enabling direct cross-modal supervision and the development of IHC-free diagnostic surrogate models. Stain-to-stain translation experiments are also supported at the semantic/patch level, though pixel-precise paired training objectives may require additional elastic registration to compensate for the serial-section deformation discussed above. This rigorous approach accounts for the fact that AMACR is a non-prostate-specific marker that can express in benign mimics, ensuring that ground truth definition was always performed strictly in the context of morphological findings alongside HMWCK basal cell staining (Ross et al., 2020). Finally, the dataset is openly available under a permissive license and accompanied by clinical metadata and preprocessing code, aligning with the FAIR principles and supporting transparent and reproducible research (Cobo et al., 2026).

### Known limitations

The dataset was collected at a single institution and includes a limited number of patients (25), which may impact the generalizability of AI models trained exclusively on these data to other scanners, staining protocols, and patient populations (Cobo et al., 2023). WSI appearance is sensitive to tissue processing, staining batch variability, and digitization equipment; models trained here may require domain adaptation before deployment in other centers. The class distribution is highly imbalanced, with tumor and stroma constituting the vast majority of annotated regions, while diagnostically critical but rare classes, such as intraductal carcinoma, atypical intraductal proliferation, and HGPIN, are represented by only a small number of instances. This imbalance poses a methodological challenge for supervised learning and should be explicitly addressed in any downstream modeling effort. Additionally, the stroma class in both modalities was generated through an automated post-processing pipeline rather than fully manual annotation, which may introduce minor boundary inaccuracies in regions of complex tissue architecture. Finally, spatial registration between H&E and IHC slides is inherently limited by tissue deformation arising from serial sectioning; while annotations have been harmonized at the semantic level, perfect pixel-to-pixel correspondence between modalities cannot be guaranteed.

### Biases and representation concerns

The dataset reflects the clinical and demographic characteristics of a single geographic region and healthcare institution in northern Spain, and therefore may not capture the full spectrum of morphological variability present across different ethnic groups, age distributions, or clinical risk profiles. Furthermore, only cases in which the final diagnosis was concordant between both uropathologists and the DeepDx Prostate (RUO) AI algorithm were included. While this criterion ensures annotation reliability, it may lead to a systematic underrepresentation of diagnostically ambiguous or discordant cases, which are particularly challenging scenarios for automated systems and those where AI assistance would provide clinically relevant information. Exact counts of screened cases excluded specifically for diagnostic discordance, as distinct from those excluded for tissue fragmentation or inadequate staining quality, were not systematically logged during retrospective curation and are therefore not reported. Accordingly, PRECISE should be understood as representing the confirmatory population of the two-stage H&E-then-IHC workflow – cases where expert and algorithmic assessments converge – rather than a population sampled to be enriched for diagnostic uncertainty. Systematic characterization of the excluded discordant cohort, and benchmarking of model performance specifically on a discordant subset, was not undertaken in the present release and is identified as a priority direction for a future extension of this resource.

### Recommendations for responsible use

This dataset supports computational pathology research and benchmarking, but models require rigorous external validation on diverse, multicenter cohorts before clinical use. Due to class imbalance, techniques like oversampling, data augmentation, or weighted loss functions should be applied to prevent bias toward majority classes. Spatial harmonization implies semantic consistency rather than pixel-level alignment; tasks requiring geometric precision may need additional image registration. Derived AI tools must act as decision-support systems alongside expert uropathologists, not as replacements. Re-identification of patients from the clinical metadata or imaging data is strictly prohibited, and any use of the dataset must comply with the terms of the associated data license and applicable research ethics standards.

## 4. Resource Availability

### 4.1 Summary Statement

PRECISE constitutes the first publicly available hybrid H&E– IHC dataset of prostate core needle biopsies with semantically harmonized, pixel-level expert annotations across both staining modalities, directly mirroring the two-stage (H&E-then-IHC) clinical diagnostic workflow used to resolve morphological uncertainty, restricted to cases in which that workflow reached diagnostic consensus. By covering the full morphological spectrum of prostate pathology — including rare and diagnostically challenging precursor lesions — it addresses a critical gap in existing public benchmarks, which focus predominantly on binary tumor classification and lack IHC pairing. This resource is designed to accelerate the development of multimodal AI systems for prostate cancer diagnosis and to promote reproducible, transparent computational pathology research.

### 4.2 Data and Code Location

The dataset is publicly available at 10.5281/zenodo.20721779. No registration or special credentials are required. To ensure reproducibility, the complete pipeline for data curation, preprocessing, and analysis is provided in the companion code repository at https://github.com/abelBEDOYA/PRECISE-data-utils. The repository includes dedicated tools for synchronized H&E and immunohistochemistry (IHC) multimodal visualization, automated morphological refinement—such as tile-based background stroma detection via morphological operations—and instance-level connected component analysis using CIELAB color space and spatial covariance features. Additionally, it features QuPath automation scripts for optimized bounding-box cropping and image pyramid compression.

### 4.3 Potential Use Cases

This dataset targets computational pathology research in the clinical domain of prostate cancer diagnosis. Primary ML tasks include multimodal semantic segmentation across paired H&E and IHC WSIs, panoptic segmentation, and self-supervised representation learning exploiting cross-stain correspondences. Secondary use cases include stain-to-stain translation, IHC-free surrogate model development, Gleason grading assistance, and domain adaptation research targeting scanner and staining variability in digital pathology.

### 4.4 Licensing

The dataset is released under the Creative Commons Attribution 4.0 International License (CC BY 4.0; https://creativecommons.org/licenses/by/4.0/). As the data were collected de novo at a single institution and are not derived from any upstream dataset, no upstream license compatibility issues apply. The accompanying code is released under the Apache License, Version 2.0 (https://www.apache.org/licenses/LICENSE-2.0).

### 4.5 Ethical Considerations

The cohort consists exclusively of adult male patients (mean age 70 years, range 49–90); no minors or vulnerable populations are included. Data were collected at the Department of Pathology, University Hospital Marqués de Valdecilla, Santander, Spain, in accordance with the Declaration of Helsinki and approved by the Cantabria Institutional Research Ethics Committee (protocol 2024.477; approved 7 February 2025). All participants provided written informed consent for data sharing and publication of anonymized microscopy images. Images and metadata were fully anonymized using SlideMaster software (3DHISTECH) prior to release, and the public release was approved jointly by the principal investigator and the IDIVAL data governance committee. For data or ethics enquiries, please contact the corresponding author.

## 5. Methods

### 5.1 Data Details

#### 5.1.1 Inclusion and Exclusion Criteria

The inclusion criteria required prostate CNBs with an established clinical indication for dual-modality histopathological evaluation. Cases were included only if there was definitive diagnostic consensus among two independent expert uropathologists and the DeepDx Prostate (RUO) AI algorithm. Given AMACR’s known non-prostate-specificity and expression in benign mimics, its interpretation was always performed in combination with HMWCK basal cell staining and in the context of morphological findings (Ross et al., 2020). Biopsies with extensive tissue fragmentation, loss of structural integrity during sectioning, or inadequate staining quality precluding definitive evaluation were excluded from the cohort.

#### 5.1.2 Data Collection Timeline

This study was conducted retrospectively using anonymized diagnostic biopsy archives. Patient selection and slide retrieval targeted cases processed between 2022 and 2024, representing real-world clinical and diagnostic pathology workflows.

#### 5.1.3 Demographic Description and Clinical Variables

As dictated by the nature of prostate pathology, the cohort consists exclusively of biologically male patients (100% male). The dataset comprises 27 WSIs corresponding to 37 prostate core needle biopsies obtained from 25 unique patients. The mean age of the cohort is 70 years (range: 49 - 90 years).

Relevant clinical and histopathological variables accompanying each case include patient age, baseline Prostate-Specific Antigen (PSA) levels, Digital Rectal Exam (DRE) findings, pelvic MRI observations, final slide-level diagnosis, ISUP Grade Group, and individual Gleason scores. These variables are systematically summarized within the structured `clinical diagnosis.txt` metadata file.

### 5.2 Methods Used for Data Creation

#### 5.2.1 Preparation, Tissue Processing and Staining Protocol

Prostate core needle biopsies were obtained via the standard transrectal approach. All biopsy specimens were fixed in 10% neutral buffered formalin, embedded in paraffin blocks, and sequentially sectioned at a thickness of 3 *μ*m.

For each selected biopsy specimen, serial sections were generated: the primary section was stained with standard Hematoxylin and Eosin (H&E), and the immediate adjacent section was subjected to double immunohistochemical (IHC) staining. The IHC process utilized a cocktail consisting of high-molecular-weight cytokeratin (HMWCK) and *α*-methylacyl-CoA racemase (AMACR), in which basal cells were visualized in brown and AMACR-expressing cells in magenta. The PIN cocktail combining HMWCK, p63 and AMACR is routinely used in clinical practice to determine the absence of basal cells as a key diagnostic criterion for prostatic adenocarcinoma (Lu et al., 2023). This double-expression and cocktail approach aligns with the established best practice guidelines for diagnostic urological immunohistochemistry (Epstein et al., 2016). The specific antibodies deployed were Monoclonal Rabbit Anti-Human AMACR (clone 13H4, ready-to-use, Dako Omnis) and Monoclonal Mouse Anti-Human High-Molecular-Weight Cytokeratin (clone 34*β*E12, ready-to-use, Dako Omnis). In this context, AMACR has been validated as a highly accurate diagnostic marker for prostate cancer on biopsy, though current evidence supports its use in combination with basal cell markers rather than as a standalone tool (Prihadi et al., 2025).

#### 5.2.2 Acquisition Equipment and Digital Pathology Protocol

A total of 27 H&E-stained slides and their corresponding 27 paired IHC-stained slides were digitized. Scanning was performed at a 20× optical magnification level using the 3DHISTECH Panoramic 250 (P250) and Panoramic 1000 (P1000) whole-slide scanners (3DHISTECH). This digitization protocol yielded high-resolution whole-slide images with a spatial resolution of 0.243 *μ*m/pixel in both the X and Y directions.

#### 5.2.3 Data Format

The original full-slide images were acquired with 3DHIS-TECH Pannoramic scanners (Pannoramic 250 for sub-01– 18; Pannoramic 1000 for sub-19–25) and are preserved in `.mrxs` format, 3DHISTECH’s native hierarchical format. From these, tissue regions containing annotations were cropped and exported as pyramidal `OME-TIFF` files with `LZW` lossless compression, comprising six resolution levels (1× to 32× downsampling) with a tile size of 512 × 512 pixels. The base-layer physical resolution is 0.243 *μ*m/pixel, and images are encoded as `RGB uint8`. Each whole-slide image has an associated semantic segmentation mask of identical dimensions and pyramid structure, ensuring exact pixel-wise correspondence between tissue appearance and ground-truth labels. Masks follow the same naming convention and `OME-TIFF` format as their source images.

The dataset consists of 25 subjects (`sub-01` to `sub-25`), each contributing at least one session of contiguous serial sections stained with hematoxylin and eosin (H&E) and HMWCK-AMACR immunohistochemistry. Accompanying metadata include `participants.csv` clinical variables (AGE, PROSTATE-SPECIFIC_ANTIGEN_(PSA)_LEVEL, DIGITAL_RECTAL_EXAM, FINDINGS_IN_PELVIC_MRI, SLIDE_DIAGNOSIS, ISUP_Grade_Group, Gleason_score, Scanner) and `label_descriptions.json`, which maps numeric labels to seven tissue classes (Tumor, Benign gland, Artifact, HGPIN, Intraductal carcinoma, Atypical intraductal proliferation, and Stroma), plus Background.

#### 5.2.4 Preprocessing and Data Anonymization Pipelines

Prior to any external storage or processing, all digital slides were rigorously anonymized using SlideMaster software (3DHISTECH) to scrub any embedded institutional headers, patient barcodes, or identifiable DICOM/WSI metadata tags. Patient identifiers were mapped to generic codes (`ANONYMOUS_CODE` and `PATIENT_NUMBER`). The tissue segmentation and patch-generation preprocessing pipelines were executed using open-source workflows. The custom preprocessing scripts are fully open-source and available in the code repository https://github.com/abelBEDOYA/PRECISE-data-utils.

#### 5.2.5 Known Artifacts and Data Limitations

Users should account for inherent histopathological and technical variation, including minor artifacts such as tissue folding, coverslip air bubbles, batch-to-batch staining intensity differences, and focal stromal retraction spaces. Because H&E and IHC images originate from physical serial sections, spatial registration is inherently limited by tissue deformation introduced during cutting and mounting; consequently, while annotations are harmonized at the semantic level, pixel-to-pixel geometric correspondence between modalities cannot be guaranteed without additional elastic registration.

### 5.3 Annotation Procedure and Ground Truth Definition

#### 5.3.1 Annotator Expertise and Consensus Workflow

The manual annotation and validation process followed a rigorous three-stage multi-observer consensus workflow to minimize diagnostic drift. The annotations were curated by two dedicated, practicing anatomical pathologists specialized in uropathology (Uropathologist A, with 8 years of experience, and Uropathologist B, with 10 years of experience).

#### 5.3.2 Ground Truth Harmonization

In the first stage, Uropathologist A independently traced and labeled all digital slides. In the second stage, Uropathologist B reviewed the entire set of annotations to score consistency, boundaries, and diagnostic agreement. Finally, both uropathologists conducted a joint consensus review on a multi-head/digital interface to resolve any boundary or diagnostic discrepancies, establishing the definitive reference standard.

Crucially, the matched CKAPM + racemase IHC slides served as a biological ground truth; in our dual immunohistochemical stain, basal cells stained brown (HMWCK-positive), whereas racemase expression (AMACR-positive) was visualized in magenta, and these markers were directly utilized by the pathologists to establish definitive pixel-level boundaries for challenging classes. In total, the dataset contains 24,387 expert-validated regional annotations covering the full morphological spectrum of prostate pathology: malignant glands, benign glands, stroma, and critical precursor or borderline entities such as HGPIN, Intraductal Carcinoma (IDC-P), and Atypical Intraductal Proliferation (AIP).

To complete the semantic maps, the Stroma class was further populated via an automated tissue-detection pipeline. Unlabelled tissue regions not covered by the manual annotations were identified through intensity thresholding in the RGB channel mean (pixels with a mean intensity below a configurable threshold, typically ~240, were classified as tissue). The resulting binary mask was refined with morphological operations (optional uniform blurring, dilation, and exterior-only erosion to smooth boundaries while preserving luminal holes) then intersected with the background class of the existing masks. Pixels satisfying both conditions were assigned to the Stroma class 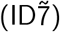. This process operates tile-by-tile with configurable overlap to avoid edge artifacts and generates pyramidal `OME-TIFF` masks fully consistent with the original multi-resolution structure.

### 5.4 AI-Readiness statement

The dataset is ready for use in DL pipelines without additional manual preprocessing. All WSIs have been digitized, anonymized, and exported in pyramidal OME-TIFF format, enabling efficient multi-resolution patch extraction via standard libraries such as TIFFFile and Zarr. Semantic segmentation masks are provided in pyramidal OME-TIFF format, with exact pixel-wise correspondence to their source images with identical dimensions, pyramid structure, and physical resolution. Each mask is a single-channel uint8 array where each pixel holds an integer class ID (0–7), directly consumable by deep learning frameworks including PyTorch and TensorFlow without reformatting. No additional label harmonization, tissue-detection thresholding, or stain normalization is strictly required prior to model training, although stain normalization may be beneficial for domain adaptation experiments given the single-center origin of the data.

## 6. Validation

### 6.1 Case Selection and Annotation Workflow

The biopsies included in this dataset were derived from a larger study designed to validate AI algorithms for the histopathological diagnosis of prostate cancer. To establish a rigorous cohort of both benign and malignant cases, all primary hematoxylin and eosin (H&E) sections were anonymized and filtered to include only cases with complete diagnostic agreement between two expert uropathologists and the DeepDx Prostate (RUO) AI algorithm. For these selected specimens, immediate adjacent sections were stained with a HMWCK + racemase immunohistochemical (IHC) cocktail to serve as a biological reference standard. Using these paired modalities, a structured three-stage manual annotation workflow was executed. Firstly, Uropathologist A independently traced and labeled all relevant histological structures—spanning malignant and benign glands, tissue artifacts, and critical precursor lesions such as HGPIN, AIP, and IDC-P—across both H&E and IHC slides. Secondly, Uropathologist B reviewed the complete set of annotations to assess consistency and boundary agreement; and finally, both experts conducted a joint consensus review to resolve any lingering discrepancies and establish the definitive, semantically harmonized pixel-level reference ground truth. This multi-stage validation process ensured a high level of diagnostic reliability, providing a robust reference benchmark for the development and evaluation of AI-based computational pathology methods.

### 6.2 Chromatic Discriminability Analysis

To assess the chromatic distinctiveness of the annotated classes, all instances were converted from RGB to CIELAB and the mean channel a* (redness) was computed per instance for tumour and benign glands. As shown in Figure 2 and the scatter plot in Figure 3, tumour regions exhibit higher a* values yet with notable overlap; a classifier relying solely on this channel would achieve an AUROC of 0.913. The residual overlap confirms that colour-based proxies remain insufficient without the morphological precision captured by expert annotations.

**Figure 2.**
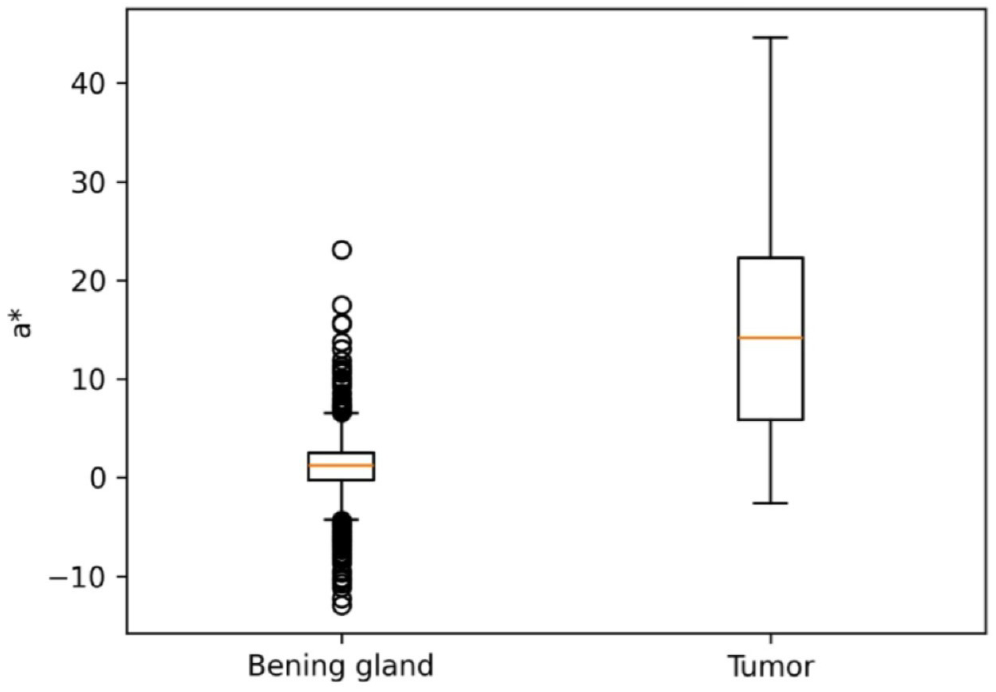
Box plot showing the distribution of the mean CIE-Lab channel a* (redness) value per annotated instance for the tumor and benign gland classes.

**Figure 3.**
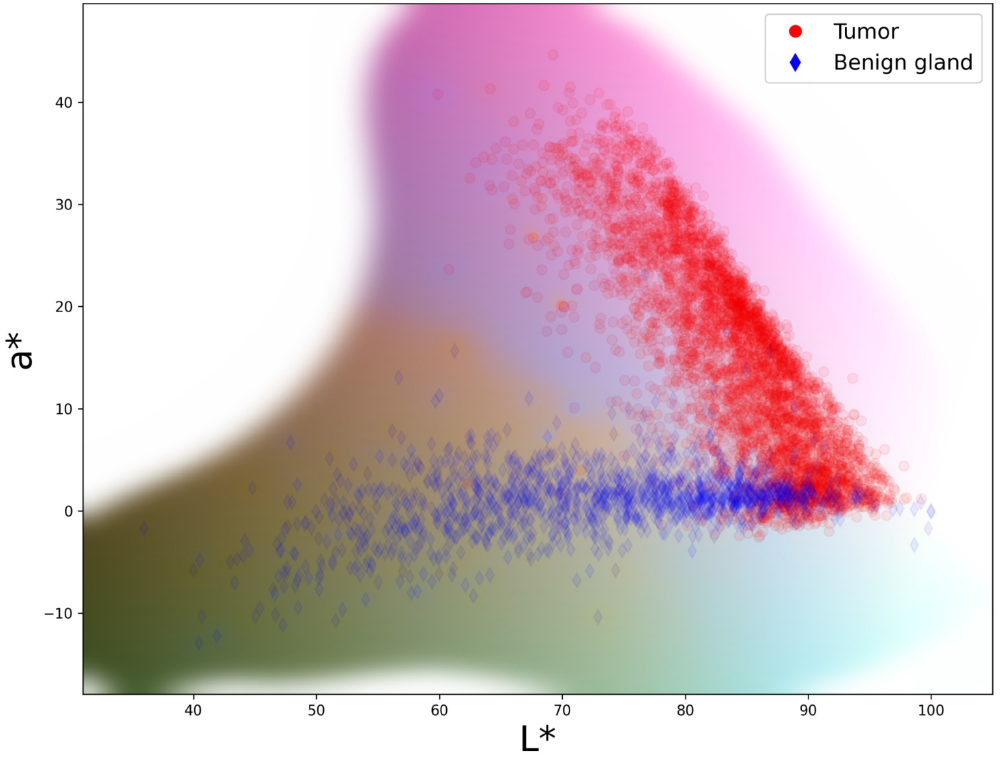
Scatter plot comparing tumor (red) and benign (blue) points in CIE-Lab space. The graph shows that while tumors tend to have a more distinct reddish profile (*a*^*\**^), there is a substantial overlap with healthy tissue, making accurate differentiation based solely on color a challenge.

## Data Availability

The dataset is publicly available at 10.5281/zenodo.20721779. No registration or special credentials are required. To en- sure reproducibility, the complete pipeline for data curation, preprocessing, and analysis is provided in the companion code repository at https://github.com/abelBEDOYA/PRECISE-data-utils.

https://zenodo.org/records/20721779

https://github.com/abelBEDOYA/PRECISE-data-utils

## 7. Acknowledgements

The authors declare that they have no competing interests.

## Notes

### Competing Interest Statement

The authors have declared no competing interest.

### Author Declarations

Cantabria Institutional Research Ethics Committee (protocol code 2024.477; approval date: 7 February 2025).

### Funding

- Universidad de Cantabria

